# A cluster randomised trial of a complex intervention to reduce anaemia and cardiometabolic risk during pregnancy and in the first year following birth in women living in rural India

**DOI:** 10.1101/2025.06.30.25330566

**Authors:** Arpita Ghosh, Aman Rastogi, Laurent Billot, Sudhir Raj Thout, Jane Hirst, D Praveen, Nicole Votruba, Sreya Majumdar

**Affiliations:** The George Institute for Global Health India, New Delhi, India; The George Institute for Global Health, Faculty of Medicine and Health, University of New South Wales, Sydney, Australia; Prasanna School of Public Health, Manipal Academy of Higher Education, India; The George Institute for Global Health, Sydney, New South Wales, Australia; Nuffield Department of Women’s and Reproductive Health, University of Oxford, Oxford, UK; The George Institute for Global Health, Imperial College London, London, UK; Oxford University Hospitals, NHS Foundation Trust, Oxford, UK; The George Institute for Global Health UK, London, UK

**Keywords:** Maternal health, Anaemia, Gestational diabetes, Preeclampsia, Clinical Trial, Cluster randomized, Statistical Analysis Plan

## Abstract

The SMART Health Pregnancy 2 trial aims to determine if the SMARThealth Pregnancy complex intervention can improve women’s health in the year after pregnancy, specifically by decreasing the prevalence of anaemia by 9% and improving screening, referral and follow-up following a pregnancy affected by either anaemia, diabetes or hypertensive disorders of pregnancy. It is designed as a parallel-group cluster randomised superiority trial. The unit of randomisation was the Primary Health Centre (PHC). A matched-pair design, where clusters are paired before randomizing one to each trial arm, was followed. This statistical analysis plan pre-specifies the method of analysis for every outcome and key variable collected in the trial. The primary outcome is the difference in the proportion of participants with any degree of anaemia (Hb measured using a point-of-care test < 12g/dL) at 12 months after delivery between the intervention and control clusters. The primary analysis will consist of a mixed effects logistic regression with anaemia at 12 months as a binary outcome, treatment group, and baseline Hb as fixed effects and PHCs as random effects. The analysis plan also includes planned sensitivity analyses including covariate adjustments and subgroup analyses.

## 2 Administrative Information

### 2.1 Study identifiers

- Protocol Version 2.4 August 2024
- Trial Registration: clinicaltrials.gov ID NCT05752955

### 2.2 Revision History

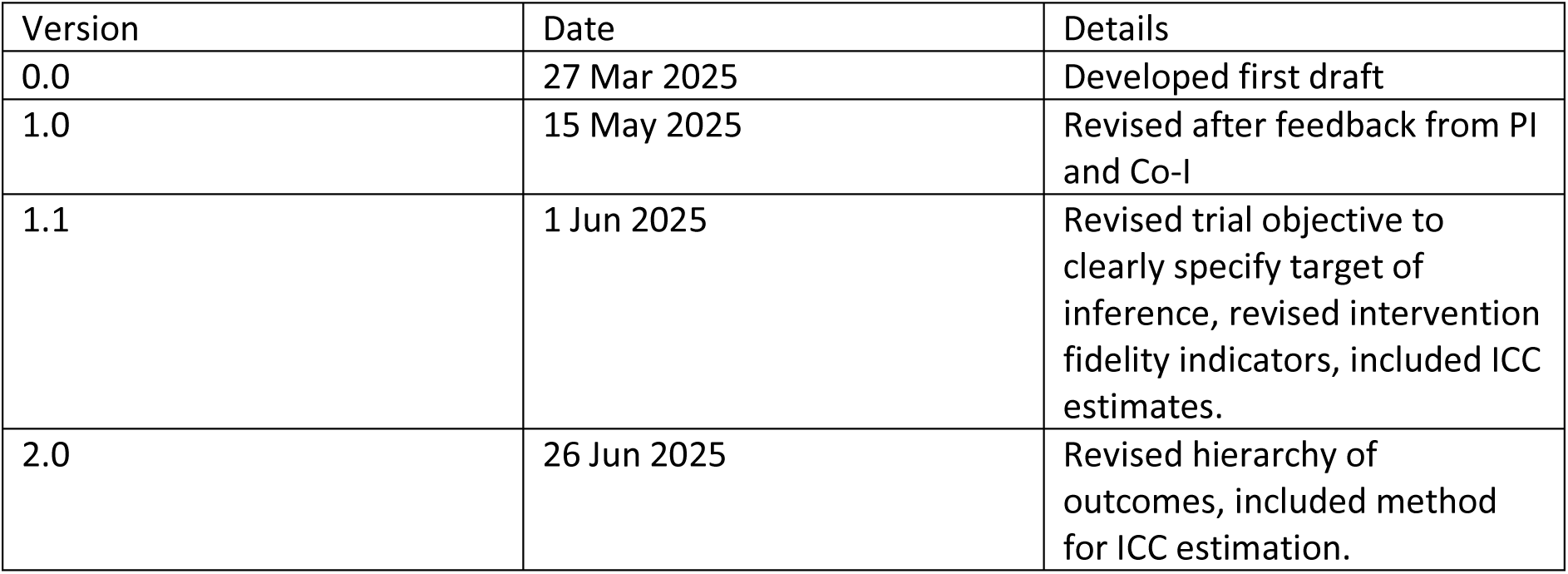

| Version | Date | Details |
| --- | --- | --- |
| 0.0 | 27 Mar 2025 | Developed first draft |
| 1.0 | 15 May 2025 | Revised after feedback from PI and Co-I |
| 1.1 | 1 Jun 2025 | Revised trial objective to clearly specify target of inference, revised intervention fidelity indicators, included ICC estimates. |
| 2.0 | 26 Jun 2025 | Revised hierarchy of outcomes, included method for ICC estimation. |

### 2.3 Contributors to Statistical Analysis Plan

| Name | Role on study | Contribution to the SAP |
| --- | --- | --- |
| Arpita Ghosh | Study statistician | Developed the SAP |
| Aman Rastogi | Study statistician | Helped develop SAP and provided critical inputs to all versions |
| Laurent Billot | Study statistician (Co-I) | Reviewed and edited every version |
| Sudhir Raj Thout | Project Manager | Implemented the project and is helping with data analysis plan |
| Jane Hirst | PI | Reviewed and provided critical input to all versions |
| D Praveen | Co-I | Reviewed and provided critical input to all versions |
| Nicole Votruba | Senior Research Fellow/Lead for Implementation analysis | Reviewed and provided critical input to all versions |
| Sreya Majumdar | Lead qualitative researcher | Reviewed and provided critical input to all versions |

## 3 Approvals

The undersigned have reviewed this plan and approve it as final. They find it to be consistent with the requirements of the protocol as it applies to their respective areas. They also find it to be compliant with ICH-E9 principles and, in particular, confirm that this analysis plan was developed in a completely blinded manner, i.e. without knowledge of the effect of the intervention(s) being assessed.

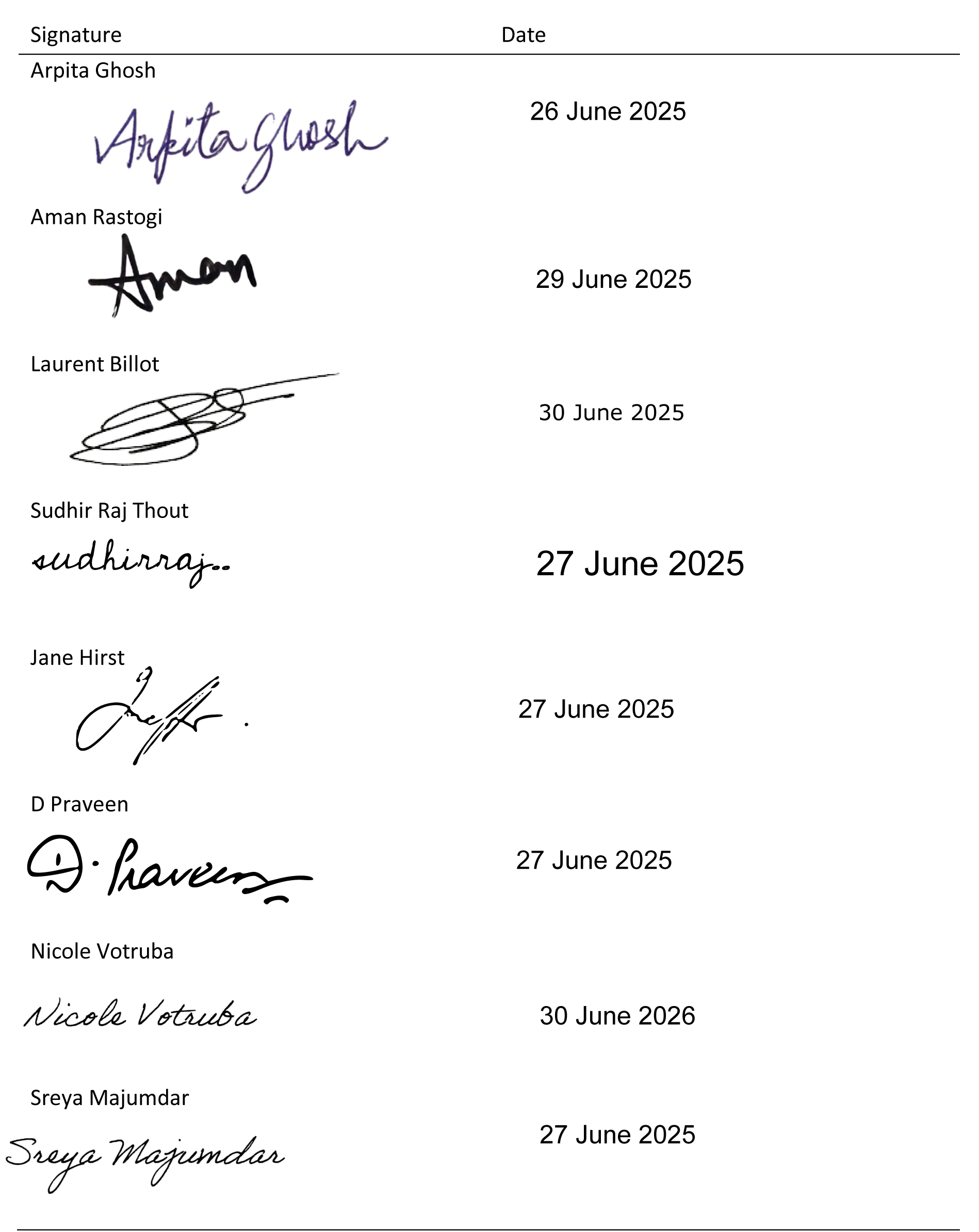

| Signature | Date |
| --- | --- |
| Arpita Ghosh | 26 June 2025 |
| Aman Rastogi | 29 June 2025 |
| Laurent Billot | 30 June 2025 |
| Sudhir Raj Thout | 27 June 2025 |
| Jane Hirst | 27 June 2025 |
| D Praveen | 27 June 2025 |
| Nicole Votruba | 30 June 2026 |
| Sreya Majumdar | 27 June 2025 |

## 4 Introduction

### 4.1 Purpose of this document

This statistical analysis plan (SAP) describes the statistical methods and data presentations to be used in the summary and analyses of data from SMARThealth Pregnancy programme. This document is based on SMARThealth Pregnancy protocol Version 2.4 dated August 2024. It describes the final analyses for evaluation of intervention at 12 months from randomization. Health economics modelling is not a part of this SAP. The document will be finalised before database lock and unblinding.

### 4.2 Study Synopsis

SMARThealth Pregnancy, adapted from Systematic Medical Appraisal, Referral and Treatment programme for cardiovascular disease prevention and management (SMARThealth), is a tool developed to improve guideline-based screening and management of anaemia, diabetes and hypertension during pregnancy and in the first year after birth. We hypothesize that the SMARThealth Pregnancy complex intervention can improve women’s health in the year after pregnancy, specifically by decreasing the prevalence of anaemia by 9% and improving screening, referral and follow-up following a pregnancy affected by either anaemia, diabetes or hypertensive disorders of pregnancy.

## 5 Study Methods

### 5.1 Trial design

This is a pragmatic, type 2 hybrid effectiveness/implementation, two-arm parallel-group cluster randomised trial involving PHCs in rural and semi-rural districts in two states in India: Siddipet district, Telangana State, and Jhajjar and Rohtak districts, Haryana State. Sixteen and fourteen PHCs in Telangana and Haryana respectively were randomised 1:1 using a matched-pair design accounting for cluster size and distance from the regional centre. Each PHC is responsible for primary care services for a population of around 30,000 people across several villages. Two villages per PHC were selected as the study sites for the trial. All community health workers and PHC staff at each study site were eligible to participate in the study. The pregnant population in the participating villages selected from participating PHC were screened to identify eligible participants. These women comprise the evaluation cohort and were followed for up to 12 months.

### 5.2 Objectives

The primary objective is to assess the effectiveness of the SMARThealth Pregnancy intervention, in women who had at least one child alive at 12 months post-delivery, in decreasing the prevalence of anaemia by 9% at 12 months after delivery. The secondary objectives are to assess the effectiveness of the intervention with respect to a range of secondary outcomes, including Hb levels, moderate to severe anaemia, women detected with GDM and/or HDP during pregnancy, and change in PHQ9, GAD7, and EQ-VAS score. This document focuses on the analyses planned to address these primary and secondary effectiveness objectives. The objective is to estimate the impact of the intervention for the average individual.

### 5.3 Eligibility

#### 5.3.1 PHC cluster eligibility

PHCs located in the selected district providing pregnancy care and the chief doctor (or equivalent) consented to participate and support the program in their centre for the duration of the study. Within each PHC, 2 villages were randomly selected to participate.

#### 5.3.2 Participant inclusion criteria

Up to 5000 pregnant women were screened until the sample size of 3240 women was reached. Potentially eligible women were identified by ANMs and ASHAs, with the support of lab technicians, at the PHC. Women were eligible if they were 12-36 weeks pregnant, as determined either by ultrasound scan, date of last menstrual period or best clinical estimate.

#### 5.3.3 Participant exclusion criteria

Women who declined participation; who plan to move away and not return to the same village in the next 12 months; women who do not speak and understand verbal communication in the local study language, or girls aged < 18 years of age.

### 5.4 Randomization

A matched-pair design, where clusters are paired before randomizing one to each trial arm, was followed. The pairing was based on:

- State (Telangana or Haryana)
- Distance in km to the district headquarters
- PHC total population size
- Local advice on population similarity (socio-economic, demographic, etc.)

Within each PHC pair, one PHC was randomly allocated to intervention using a computer-generated randomisation programme. The other PHC was then designated as the control. To ensure a 1:1 balance (the same number of PHCs allocated to intervention and to control) within each state, we selected an even number of PHCs in each state. Sixteen PHCs were enrolled from Telangana and 14 from Haryana.

### 5.5 Sample size

With 15 clusters (PHCs) in each arm, we estimated that we need 86 women per cluster (total sample size 2580) to detect a change in anaemia rates by 9% over 3 years (i.e., 53% to 44%) with 80% power at 5% level of significance, assuming an ICC of 0.02 (based on pilot data). Allowing for losses to follow-up of 20%, the total sample size needed is 3240. Pilot data showed that with 2 villages per PHC it will be feasible to recruit 108 women per PHC site in 18 months.

### 5.6 Framework

We will test for superiority of the primary and secondary outcomes.

### 5.7 Statistical interim analysis and stopping guidance

No interim analyses were conducted.

### 5.8 Timing of final analysis

Final analysis for all outcomes will be conducted after final assessments (9-12 months post-delivery) are completed.

### 5.9 Timing of outcome assessments

The following table shows the timing of the assessments that will be used to define primary and secondary outcomes.

| Outcome | Population | Mode | Baseline (12-36 weeks of gestation) | Midline (6-10 weeks post-delivery) | Final (9-12 months post-delivery) |
| --- | --- | --- | --- | --- | --- |
| Hb measured using a point-of-care test | ALL women | Measurement | X |  | X |
| Blood pressure and pulse | ALL women | Measurement | X |  | X |
| EQ5D | ALL women | Questionnaire | x |  | X |
| PHQ9 | ALL women | Questionnaire | X |  | X |
| GAD7 | ALL women | Questionnaire | X |  | X |
| Diagnosed with GDM during pregnancy | ALL women | Questionnaire |  | X | X |
| Diagnosed with HDP during pregnancy | ALL women | Questionnaire |  | X | X |
| Participants referred to a specialist because of a high-risk pregnancy condition | Women who had any high-risk condition diagnosed during pregnancy (defined based on baseline, midline and endline assessments) | Questionnaire |  | X | X |
| Had glucose/bp assessment after delivery | Women who reported being diagnosed with GDM/HDP during midline & endline assessments | Telephonic interviews of women after all final assessments were over |  |  |  |
| Diagnosed with diabetes after delivery | Women detected with GDM | Questionnaire |  | X | X |
| Diagnosed with high bp after delivery | Women detected with HDP | Questionnaire |  | X | X |
| IYCF indicators | All live births | Questionnaire |  |  | X |

## 6 Statistical Principles

### 6.1 Confidence intervals and P values

All statistical tests will be 2-sided and will be performed using a 5% significance level. No multiplicity adjustment will be made as there is only one primary outcome and the secondary outcomes are for supporting evidence.

### 6.2 Adherence and Protocol deviations

In the intervention arm, women will be expected to have 2 additional ANC visits and 5 PNC visits as part of the SMARThealth Pregnancy intervention. At each visit, haemoglobin and blood pressure will be measured. The electronic decision support system includes risk assessment and recommendation for PHC doctor review.

These components will be assessed in the participants in the intervention arm using the following indicators

1. % of participants who received all 7 ANC/PNC visits
2. % of participants who received 5 PNC visits
3. Mean number of ANC/PNC visits
4. Mean number of in-person ANC/PNC visits
5. % participants who had bp assessed
6. Mean number of bp assessments
7. % participants who had capillary Hb checked during pregnancy
8. % participants who had capillary Hb checked post-delivery
9. Mean number of Hb assessments in cluster
10. % participants with a high-risk condition (high bp or anaemia) who had doctor visit

These cluster-level values will be summarised using median (IQR).

### 6.3 Analyses populations

All outcomes will be analysed using a modified intention-to-treat (mITT) population. The ITT population includes all women who were randomized. The mITT population will exclude women who had an abortion or miscarriage or still birth or if the child died after birth as these women were not interviewed in anticipation that that they most likely will decline ongoing participation in the study. However, for multiple pregnancies, women with at least one child alive were followed up. Characteristics and outcomes assessed at baseline for women with adverse birth outcomes will be reported separately. The mITT population will be analysed according to the group they were randomised to and regardless of whether they received the intervention as intended or not. There will be women who will be excluded from analyses due to missing outcome data. This may be explored in sensitivity analyses e.g. imputations (see Section 8.2.1.4 for details about missing data handling).

‘Per-protocol’ and ‘as-treated’ analyses will not be part of main analyses and will be conducted post-hoc as secondary analyses/separate publications.

## 7 Trial Population

### 7.1 Recruitment

CONSORT diagram comprising the number of clusters and participants assessed for eligibility, randomised, followed-up and analysed will be presented.

### 7.2 Withdrawal/lost to follow-up

All withdrawals/lost to follow-up will be listed.

### 7.3 Baseline participant characteristics

Participants will be described with respect to their background characteristics such as age, height, weight, religion, marital status, diet, substance use, education, income, employment, past medical history, past and current pregnancy details, family history, clinical biomarkers (haemoglobin and blood pressure), quality of life, PHQ-9 and GAD-7 scores. Categorical data will be summarised by numbers and percentages. Continuous data will be summarised by mean, SD, median, IQR, and range. Baseline characteristics will be summarized by treatment group for the ITT and mITT populations. Tests of statistical significance will not be performed for baseline characteristics; rather, the clinical importance of any imbalance will be noted.

Cluster characteristics such as PHC total population size, distance from district headquarters, PHC staffing and birth rate will be presented for the two arms.

## 8 Analysis

### 8.1 Outcome definitions

Participant-level outcomes of the trial are:

#### Primary

The difference in the proportion of participants with any degree of anaemia (Hb measured using a point-of-care test < 12g/dL) at 12 months after delivery between the intervention and EUC clusters

#### Secondary

1. Difference in the proportion of participants in different categories of anaemia: no anaemia > 12.0 g/dL; mild anaemia 11.0–11.9 g/dL; moderate anaemia 8.0–10.9 g/dL; severe anaemia < 8 g/dL (per WHO thresholds used for non-pregnant adults) at 12 months after delivery between the intervention and EUC clusters
2. Mean difference in Hb measured using a point-of-care test at 12 months after delivery between the intervention and EUC clusters.
3. Difference in proportion of women detected with GDM and/or HDP during pregnancy, between the intervention and EUC arms
4. Difference between the intervention and EUC arms in change between baseline and 12 months in PHQ9, GAD7 and EQ-VAS score

#### Exploratory

1. Among women who were diagnosed with a high-risk condition during pregnancy, the difference in proportion of women who were referred to a specialist because of the high-risk condition, between the intervention and EUC arms
2. Among women detected with GDM and/or HDP during pregnancy, the difference in proportion of women who underwent glucose/bp assessments after delivery (glucose assessment for those with GDM and BP assessment for those with HDP), between the intervention and EUC arms
3. Among women detected with GDM and/or HDP during pregnancy, the difference in proportion of women who were diagnosed with diabetes (those with GDM) and high bp (those with HDP) after delivery, between the intervention and EUC arms
4. Among women detected with HDP during pregnancy, the difference in change between baseline and 12 months in blood pressure (systolic, diastolic and mean arterial pressure), between the intervention and EUC arms
5. Difference between the intervention and EUC arms in WHO Infant and Young Child Feeding indicators

1. Proportion of children who were ever breastfed
2. Proportion of children who had early initiation of breastfeeding (put to the breast within 1 hour of birth)
3. Proportion of children who were fed exclusively with breast milk for the first two days after birth
4. Proportion of children who were who were fed breast milk during the previous day
5. Proportion of children who were fed from a bottle with a nipple during the previous day
6. Proportion of children who consumed a sweet beverage during the previous day

### 8.2 Analysis methods

#### 8.2.1 Analyses of primary outcome

##### 8.2.1.1 Main analysis

Anaemia at 12 months after delivery will be defined as Hb <12g/dL. It will be analysed using mixed effects logistic regression with anaemia at 12 months as a binary outcome, treatment group, and baseline Hb as fixed effects and PHCs as random effects. Even though we used a matched-pair design to randomize the clusters, we will not adjust for matching since this has been shown to provide unbiased estimates of the intervention effect and provide similar power to a matched analysis.^1^ The effect of the intervention will be estimated as the odds ratio (OR) together with its 95% confidence interval. This model will give valid estimates in case of data missing at random (see Section 8.2.1.4 for details about missing data handling). Risk difference along with 95% CI will be estimated using a linear model (i.e. assuming a normal distribution and identity link).^2^

##### 8.2.1.2 Adjusted analyses

The analysis described above will be rerun after adding the following baseline participant-level covariates: age (continuous), bmi (continuous), number of previous pregnancies, high risk pregnancy (yes/no), education (none, primary, secondary, more than secondary). Subsequently, another model will be run by adding few PHC-level covariates – total population size, distance from district headquarters, staffing and birth rate, to the above model. Adjusted analyses will be performed as a sensitivity analysis and the adjusted treatment effect will be reported as adjusted OR and 95% CI.

##### 8.2.1.3 Subgroup analyses

Subgroup analyses will be conducted according to the following subgroups:

- Site (Haryana vs. Telangana)
- PHC total population size categorized using median
- Distance from district headquarter categorized using median
- Age group: < 30 years vs. ≥ 30 years
- Number of previous pregnancies: 0 vs. 1 or more
- High risk pregnancy (yes/no)

Analyses for each subgroup will be performed by adding the subgroup variable and its interaction with the treatment group as fixed effects to the model described in section 8.2.1.1. For each subgroup, odds ratio and 95% confidence interval will be presented. The results will be displayed on a forest plot including the p-value for heterogeneity corresponding to the interaction term between the intervention and the subgroup variable.

##### 8.2.1.4 Missing data

The main analysis makes valid inference under the missing at random (MAR) assumption. A tipping point sensitivity analysis will be performed using the approach described by Cro et al^3^ to assess the effect of missing data under the assumption that data are not missing at random (NMAR).

As a starting point, we will first run an imputation model under the missing at random (MAR) assumption. This MAR imputation model will use fully conditional specification (FCS)^4, 5^ and will include the following variables: Hb at baseline and 12 months, a variable indicating the PHC cluster, state, a variable indicating the treatment group, and all key sociodemographic, clinical and medical baseline variables (e.g. age, education, household income, height, weight, sbp, dbp, number of previous pregnancies, high risk pregnancy). Hb and other missing continuous variables will be imputed using linear regression while categorical variables will be imputed using either a logistic model (for binary variables) or a discriminant function method (for nominal variables). One hundred sets of imputed data will be created.

In tipping point analysis, the MI imputed values will be adjusted using a pair of shift parameters, one for each arm. The shift parameters will vary across a range of values, allowing for scenarios where subjects missing values in one arm may have worse outcomes than subjects missing values in the other arm. For each pair of pre-specified shift parameters, the MI imputed values will be adjusted using this pair and we will have 100 modified-imputed datasets. The model specified in section will be fit to each of the 100 modified-imputed datasets and the results will be pooled across the 100 datasets using Rubin’s rule. The shift parameters will be set in incremental sequences usually in the direction to reduce the treatment effect until statistical significance is lost, i.e. two-sided p-value is >0.05.

Additional imputation analyses will be considered post-hoc.

#### 8.2.2 Analyses of secondary outcomes

##### 8.2.2.1 Main analyses

###### 8.2.2.1.1 Proportion of participants with no, mild, moderate and severe anaemia

This will be analysed as an ordinal variable with 4 levels using ordinal logistic regression with treatment group and baseline Hb as fixed effects and PHC as a random effect. The intervention effect will be estimated as the OR of higher level of anaemia and its 95% CI using the control arm as the reference.

We will test the proportional-odds assumption using Brant test.^6^ In case of violation, we will still proceed with the analysis and interpret the intervention OR as an average effect across all anaemia categories but with the understanding that it may not be constant across all levels. This will be complemented by a graphical assessment of shifts across categories as well as a binary analysis (see Section 8.2.4.1). As a sensitivity analysis, we will apply partial proportional odds logistic regression,^7^ relaxing the proportional odds assumption for covariates where it does not hold.

###### 8.2.2.1.2 Mean difference in Hb measured using a point-of-care test at 12 months after delivery between the intervention and EUC clusters

Mean difference in Hb at 12 months after delivery will be analysed using a linear mixed effects model with treatment group, and baseline Hb as fixed effects and PHC as a random effect. The effect of the intervention will be estimated as the adjusted mean difference at 12 months together with its 95% confidence interval.

###### 8.2.2.1.3 Difference in proportion of women detected with GDM and/or HDP during pregnancy, between the intervention and EUC arms

This binary outcome will be analysed using mixed effects logistic regression with treatment group as a fixed effect and PHC as a random effect in the model. The effect of the intervention will be estimated as the odds ratio (OR) together with its 95% confidence interval. If there is convergence issue, the PHC random effect will be dropped.

###### 8.2.2.1.4 Difference between the intervention and EUC arms in change between baseline and 12 months in PHQ9, GAD7 and EQ-VAS score

The secondary outcomes - change from baseline in EQ-VAS score, GAD-7, and PHQ-9 score at 12 months, will be analysed using the same approach as above - a linear mixed model with treatment group, and baseline score as fixed effects and PHC as a random effect. The effect of the intervention will be estimated as the mean difference in change from baseline at 12 months together with its 95% confidence interval.

##### 8.2.2.2 Adjusted analyses

The analyses described above will be rerun after adding the covariates specified for the primary outcome in Section 8.2.2.18.2.1.2. Adjusted analyses will be performed as a sensitivity analysis and the adjusted treatment effect will be reported either as adjusted mean difference or adjusted OR and its 95% CI.

#### 8.2.3 Analyses of exploratory outcomes

##### 8.2.3.1 Among women who were diagnosed with a high-risk condition during pregnancy, the difference in proportion of women who were referred to a specialist because of the high-risk condition, between the intervention and EUC arms

This binary outcome will be analysed using mixed effects logistic regression with treatment group as a fixed effect and PHC as a random effect in the model. The effect of the intervention will be estimated as the odds ratio (OR) together with its 95% confidence interval. If there is convergence issue, the PHC random effect will be dropped.

##### 8.2.3.2 Among women who reported being diagnosed with GDM and/or HDP during pregnancy at the time of midline assessment, the difference in proportion of women who underwent glucose assessments (those with GDM) and bp assessments (those with HDP) after delivery, between the intervention and EUC arms

##### 8.2.3.3 Among women detected with GDM and/or HDP during pregnancy, the difference in proportion of women diagnosed with diabetes (those with GDM) and high bp (those with HDP) after delivery, between the intervention and EUC arms

Given the small number of women, these will be examined descriptively.

##### 8.2.3.4 Among participants detected with HDP during pregnancy, mean difference in change between baseline and 12 months in systolic bp, diastolic bp and mean arterial pressure, between the intervention and EUC arms

The secondary outcomes - change from baseline in systolic bp, diastolic bp and mean arterial pressure in participants detected with HDP during pregnancy, will be analysed using a linear mixed model with treatment group and baseline systolic bp, diastolic bp and mean arterial pressure, respectively, as fixed effects and PHC as a random effect. The effect of the intervention will be estimated as the mean difference in change from baseline at 12 months together with its 95% confidence interval. If there is convergence issue, the PHC random effect will be dropped.

##### 8.2.3.5 Difference between the intervention and EUC arms in WHO Infant and Young Child Feeding indicators

These binary outcome variables will be analyzed using the same approach as in 8.2.1.1.

#### 8.2.4 Additional analyses

##### 8.2.4.1 Binary analyses of anaemia

A binary analysis will be performed by considering participants with moderate/severe anaemia (Hb < 11 g/dL) vs. no/mild anaemia (Hb ≥ 11 g/dL) at 12 months after delivery between the intervention and EUC clusters. This analysis will be conducted using the same approach as for the primary outcome – a mixed effects logistic regression with treatment group and baseline Hb as fixed effects and PHCs as random effects. The effect of the intervention will be presented as the OR of a poor outcome with associated 95% CI. A similar analysis will be performed on severe anaemia alone (Hb < 8 g/dL).

##### 8.2.4.2 Disaggregated analyses by geography

All analyses will be carried out separately by geography (Haryana and Telangana) and will be reported as supplementary material.

##### 8.2.4.3 ICC estimates

ICC estimates for the primary and secondary outcomes will be reported. Estimates at baseline and at endline separately for the intervention and control arms will be reported. These will be estimated from the mixed effects model used in main analysis assuming a nested/block exchangeable covariance structure.^8^

## Data Availability

This manuscript is a statistical analysis plan for a trial. The corresponding trial is ongoing and data are not yet available.

## Appendix: Proposed tables and figures

**Table 1.**
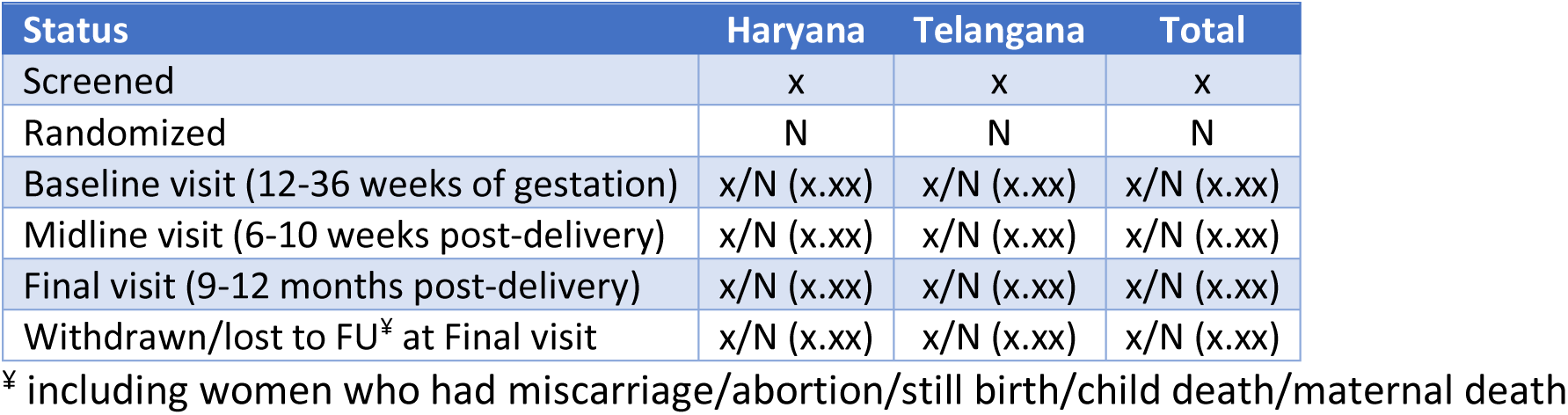
Subject disposition.

**Table 2.** Baseline characteristics of study participants.

|  | ITT population<sup>‡</sup> |  | mITT population<sup>¶</sup> |
| --- | --- | --- | --- |
| Characteristic | Intervention<br>N= | Control<br>N= |  |
| **Age (years)** |  |  |  |
| N |  |  |  |
| Mean (SD) |  |  |  |
| Median (IQR) |  |  |  |
| Min, Max |  |  |  |
| **Height (cm)** |  |  |  |
| N |  |  |  |
| Mean (SD) |  |  |  |
| Median (IQR) |  |  |  |
| Min, Max |  |  |  |
| **Weight (kg)** |  |  |  |
| N |  |  |  |
| Mean (SD) |  |  |  |
| Median (IQR) |  |  |  |
| Min, Max |  |  |  |
| **BMI (kg/m2)** |  |  |  |
| N |  |  |  |
| Mean (SD) |  |  |  |
| Median (IQR) |  |  |  |
| Min, Max |  |  |  |
| **Religion** |  |  |  |
| Hindu |  |  |  |
| Others<sup>§</sup> |  |  |  |
| **Participant is married** |  |  |  |
| Yes |  |  |  |
| No |  |  |  |
| **Diet** |  |  |  |
| Vegetarian/Vegan |  |  |  |
| Others<sup>f</sup> |  |  |  |
| **Education** |  |  |  |
| Illiterate |  |  |  |
| Upto secondary school |  |  |  |
| Under graduation and above<sup>g</sup> |  |  |  |
| **Substance use** |  |  |  |
| Cigarettes |  |  |  |
| Paan |  |  |  |
| Alcohol |  |  |  |
| **Self employment** |  |  |  |
| None<sup>h</sup> |  |  |  |
| Part time |  |  |  |
| Full time |  |  |  |
| **Husband's employment** |  |  |  |
| None<sup>h</sup> |  |  |  |
| Part time |  |  |  |
| Full time |  |  |  |
| **Household income** |  |  |  |
| N |  |  |  |
| Mean (SD) |  |  |  |
| Median (IQR) |  |  |  |
| Min, Max |  |  |  |
| **Gestational age (weeks)** |  |  |  |
| N |  |  |  |
| Mean (SD) |  |  |  |
| Median (IQR) |  |  |  |
| Min, Max |  |  |  |
| **No. of previous pregnancies** |  |  |  |
| N |  |  |  |
| Mean (SD) |  |  |  |
| Median (IQR) |  |  |  |
| Min, Max |  |  |  |
| **No. of live births** |  |  |  |
| N |  |  |  |
| Mean (SD) |  |  |  |
| Median (IQR) |  |  |  |
| Min, Max |  |  |  |
| **Any high-risk condition in previous pregnancies** |  |  |  |
| Yes |  |  |  |
| No |  |  |  |
| **Any high-risk condition diagnosed in current pregnancy before baseline visit** |  |  |  |
| Yes |  |  |  |
| **No** |  |  |  |
| **Diagnosed with any disease such as cardiac disease, respiratory disease, kidney disease, malaria, TB and so on** |  |  |  |
| **Yes** |  |  |  |
| **No** |  |  |  |
| **Family history of heart disease/diabetes/high blood pressure/stroke** |  |  |  |
| **Yes** |  |  |  |
| **No** |  |  |  |
| **Take any regular medications** |  |  |  |
| **Yes** |  |  |  |
| **No** |  |  |  |
| **Systolic blood pressure (mmHg)** |  |  |  |
| **N** |  |  |  |
| **Mean (SD)** |  |  |  |
| **Median (IQR)** |  |  |  |
| **Min, Max** |  |  |  |
| **Diastolic blood pressure (mmHg)** |  |  |  |
| **N** |  |  |  |
| **Mean (SD)** |  |  |  |
| **Median (IQR)** |  |  |  |
| **Min, Max** |  |  |  |
| **Mean arterial pressure** |  |  |  |
| **N** |  |  |  |
| **Mean (SD)** |  |  |  |
| **Median (IQR)** |  |  |  |
| **Min, Max** |  |  |  |
| **Pulse (beats per minute)** |  |  |  |
| **N** |  |  |  |
| **Mean (SD)** |  |  |  |
| **Median (IQR)** |  |  |  |
| **Min, Max** |  |  |  |
| **Haemoglobin (g/dL)** |  |  |  |
| **N** |  |  |  |
| **Mean (SD)** |  |  |  |
| **Median (IQR)** |  |  |  |
| **Min, Max** |  |  |  |
| **EQ-VAS score** |  |  |  |
| **N** |  |  |  |
| **Mean (SD)** |  |  |  |
| Median (IQR) |  |  |  |
| Min, Max |  |  |  |
| **PHQ-9 score** |  |  |  |
| N |  |  |  |
| Mean (SD) |  |  |  |
| Median (IQR) |  |  |  |
| Min, Max |  |  |  |
| **GAD-7 score** |  |  |  |
| N |  |  |  |
| Mean (SD) |  |  |  |
| Median (IQR) |  |  |  |
| Min, Max |  |  |  |

<sup>u</sup> All women who were randomized
<sup>¶</sup> Excluding women who had miscarriage/abortion/still birth/child death after birth from ITT population

**Table 3.** Characteristics of PHCs.

| Characteristic | Intervention<br>(n=15) | Control<br>(n=15) |
| --- | --- | --- |
| **Total population size** |  |  |
| Mean (SD) |  |  |
| Median (Q1,Q3) |  |  |
| Min-Max |  |  |
| **Distance from district headquarters** |  |  |
| Mean (SD) |  |  |
| Median (Q1,Q3) |  |  |
| Min-Max |  |  |
| **Staffing** |  |  |
| Mean (SD) |  |  |
| Median (Q1,Q3) |  |  |
| Min-Max |  |  |
| **Births/birth rate** |  |  |
| Mean (SD) |  |  |
| Median (Q1,Q3) |  |  |
| Min-Max |  |  |

**Table 4.** Intervention fidelity.

| Characteristic of PHC cluster in Intervention arm | Median (IQI) |
| --- | --- |
| % of participants who received all 7 ANC/PNC visits |  |
| % of participants who received 5 PNC visits |  |
| Mean number of ANC/PNC visits |  |
| Mean number of in-person ANC/PNC visits |  |
| Proportion of ANC/PNC visits that are in-person |  |
| % participants who had bp assessed |  |
| Mean number of bp assessments |  |
| % participants who had capillary Hb checked during pregnancy |  |
| % participants who had capillary Hb checked post-delivery |  |
| Mean number of Hb assessments |  |
| % participants with a high-risk condition (high bp or anaemia) who were referred to doctor |  |
| % participants with a high-risk condition (high bp or anaemia) who had doctor visit |  |

**Table 5.**
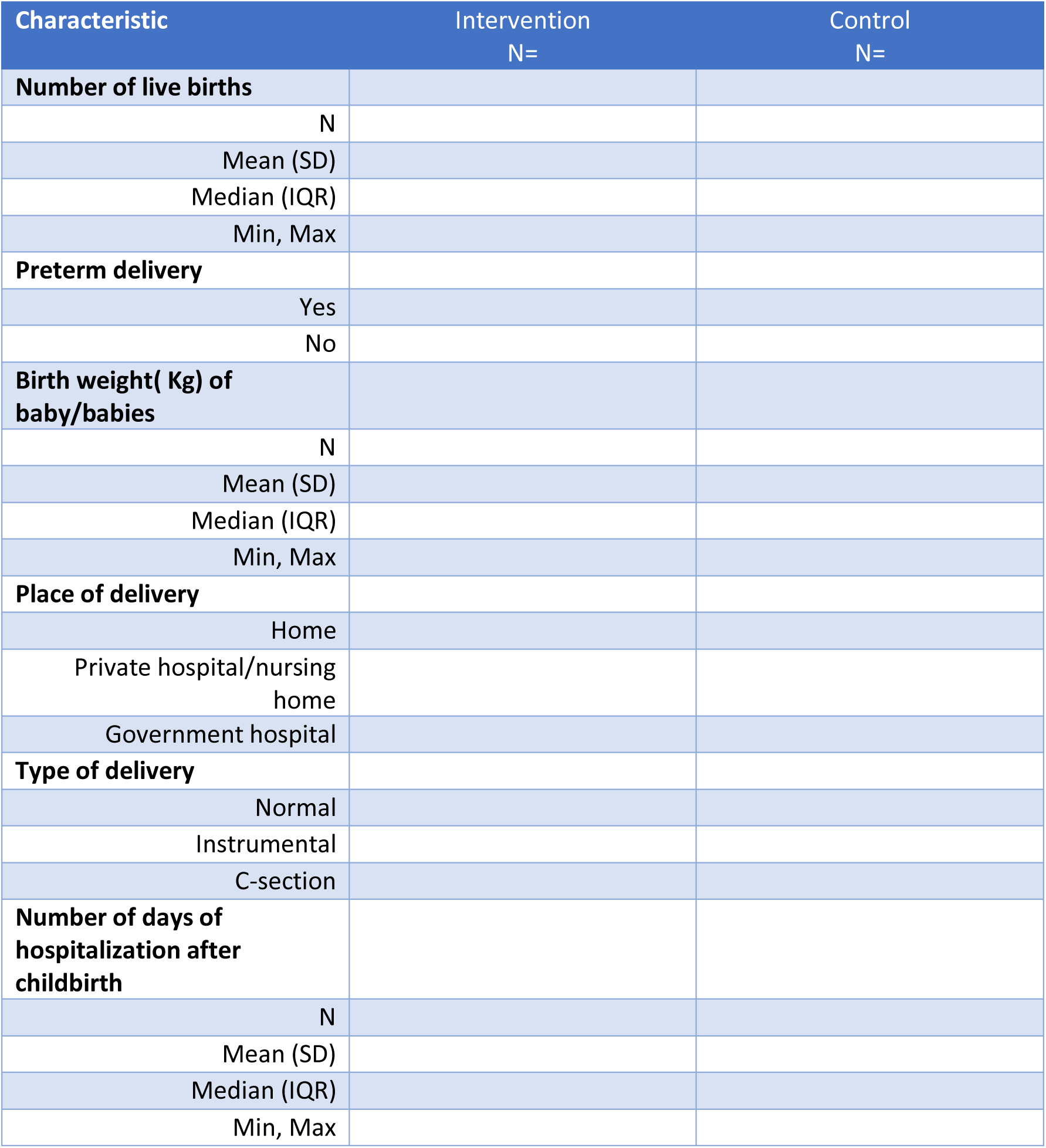
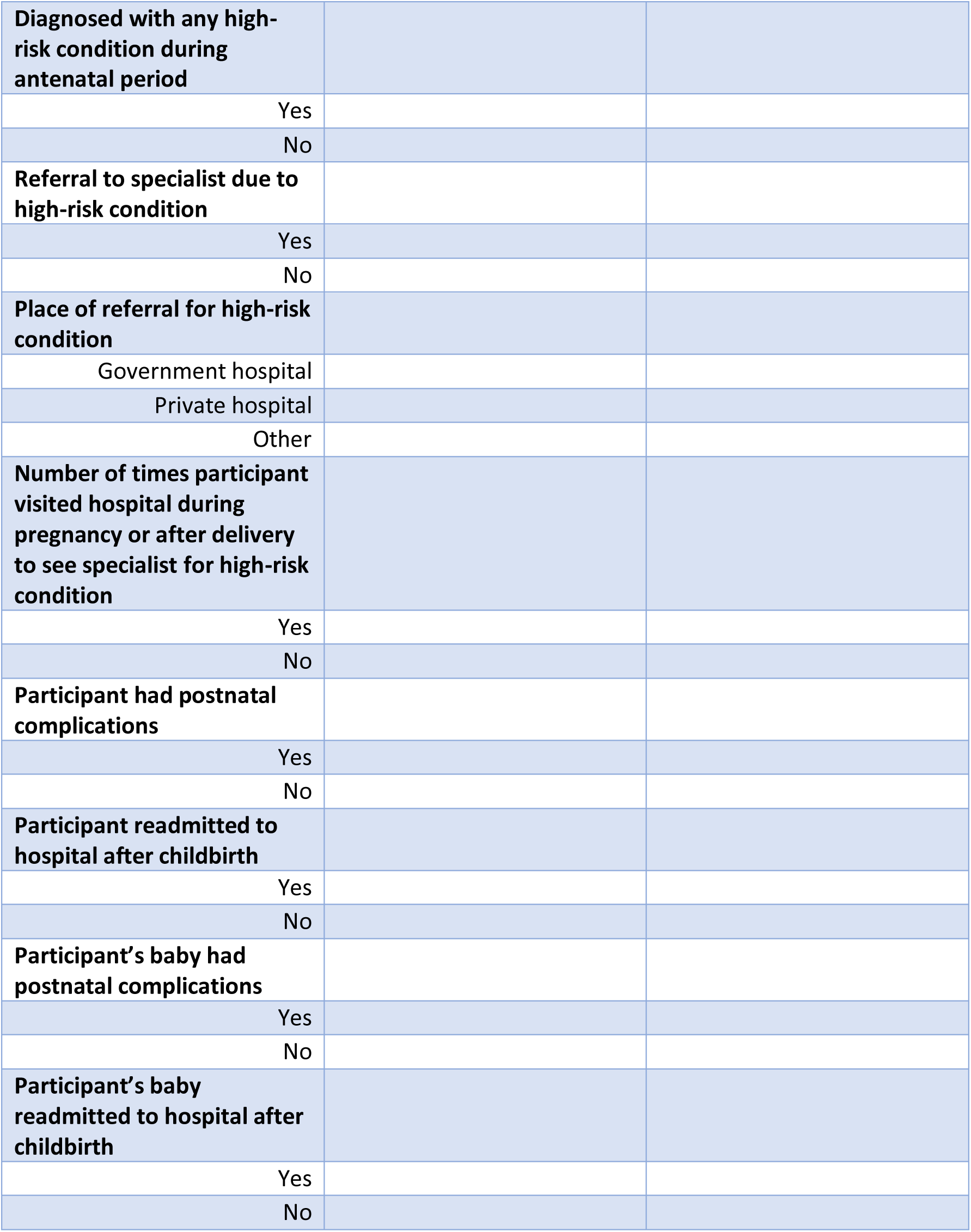
Maternal and pregnancy outcomes.

**Table 6.**
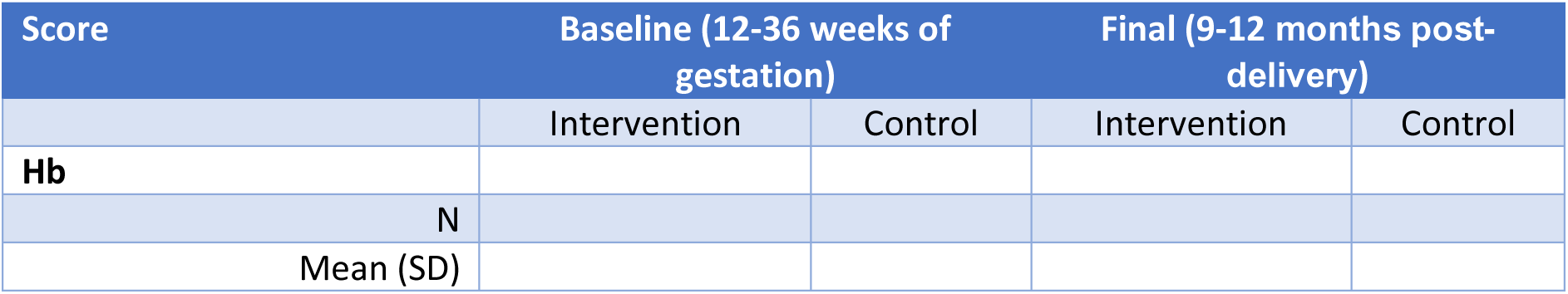
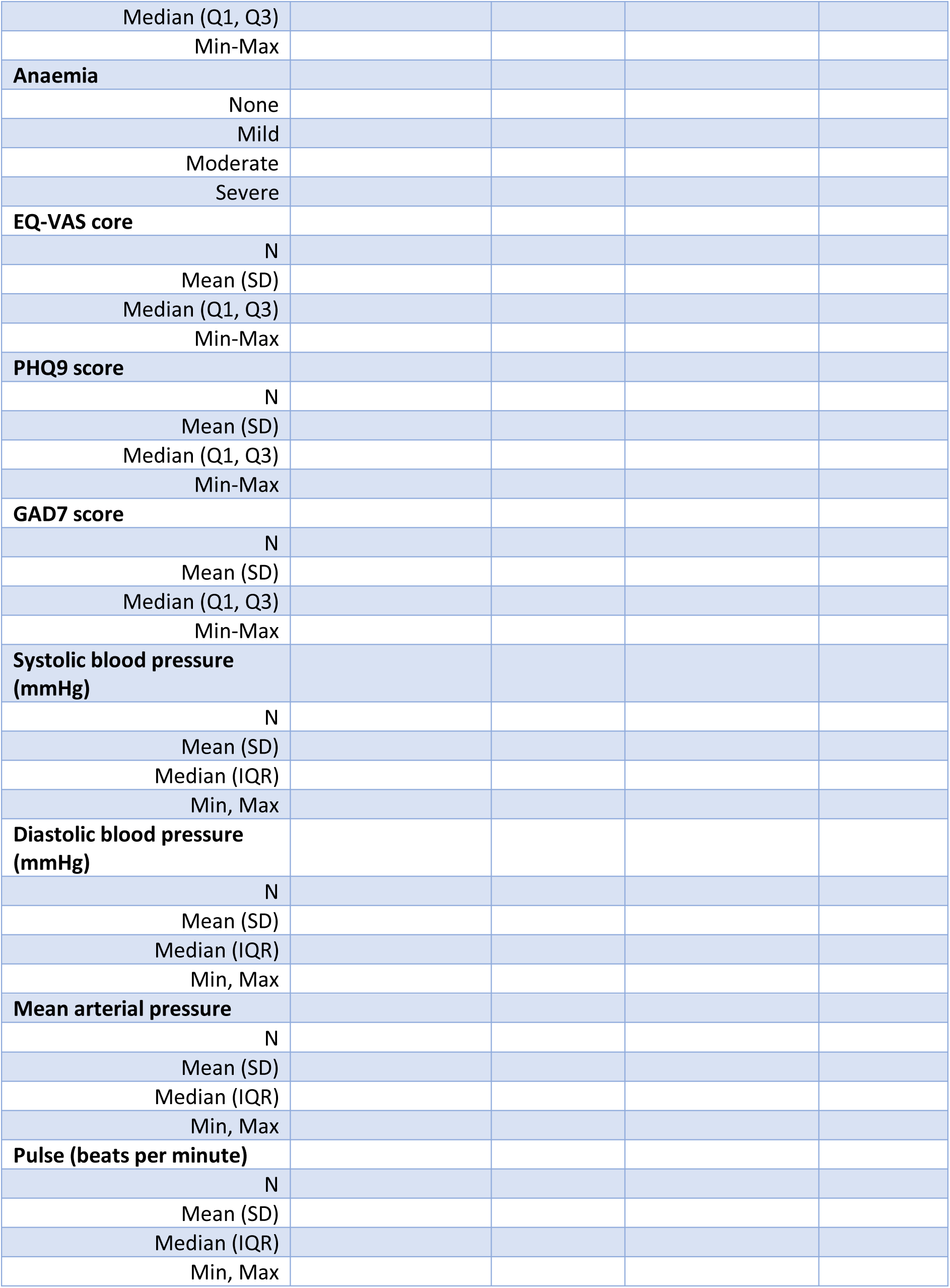
Summary of outcome measurements at baseline and final visits, by treatment arm.

**Table 7.**
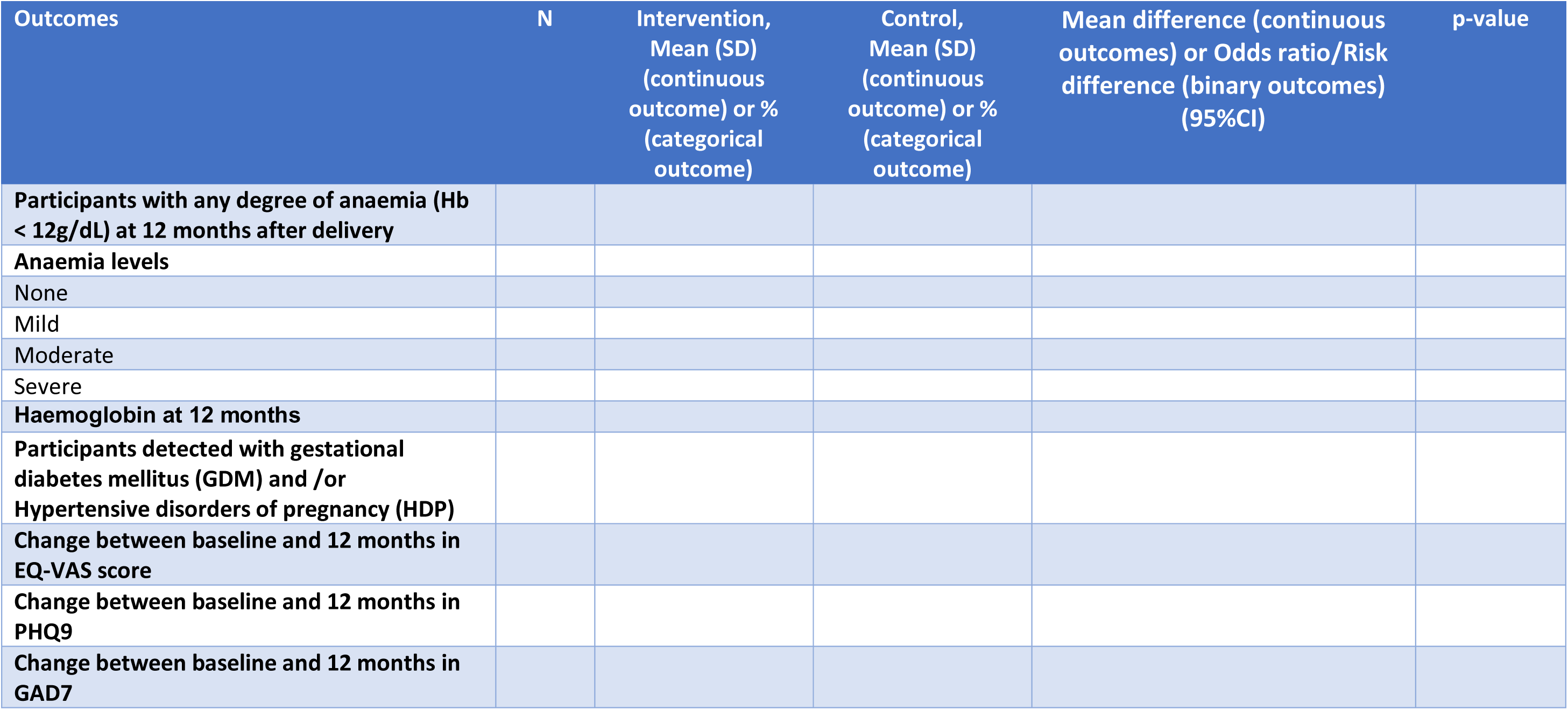
Analysis of primary and secondary outcomes.

**Table 8.**
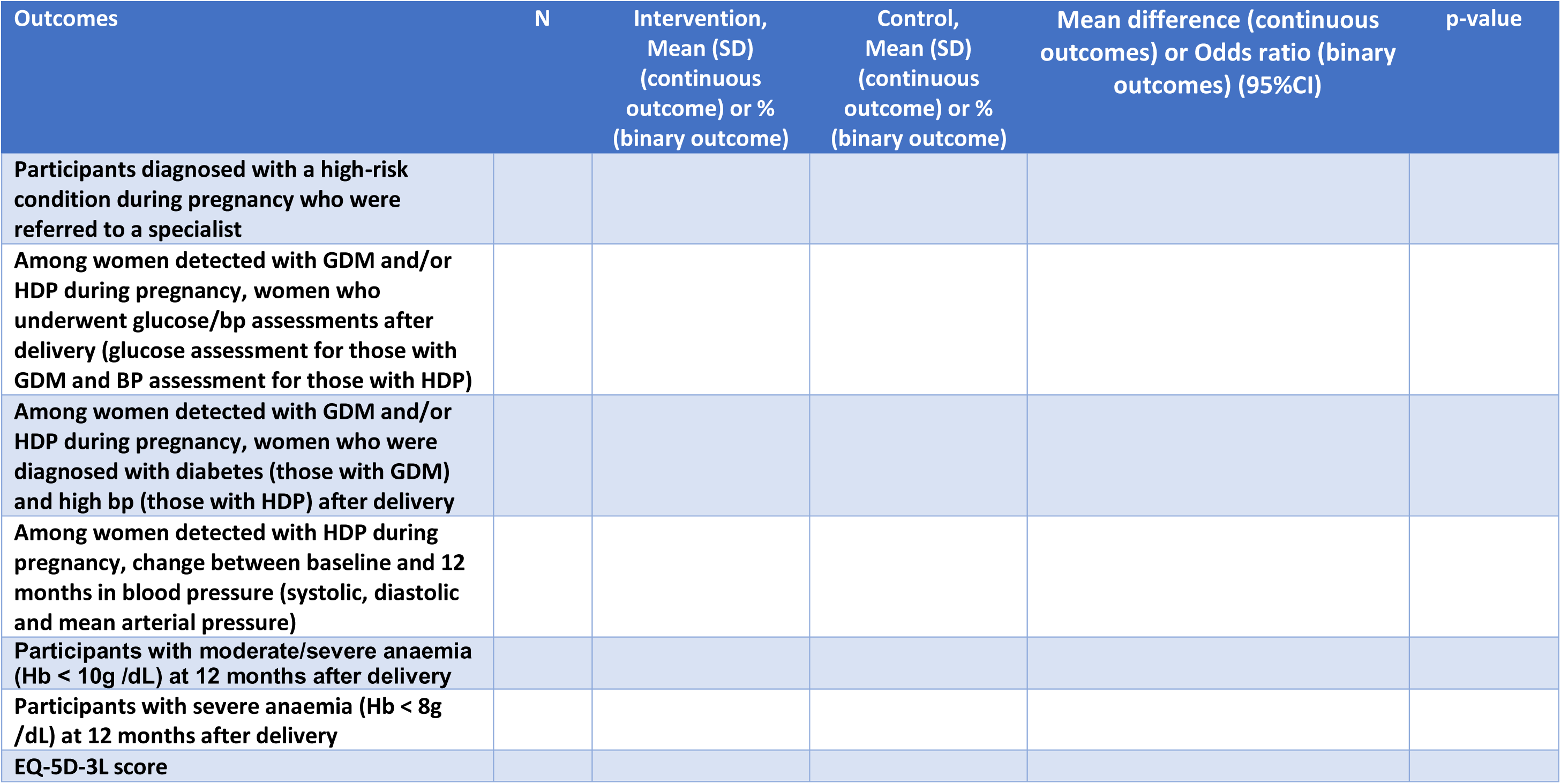
Analyses of exploratory and additional outcomes.

**Table 9.**
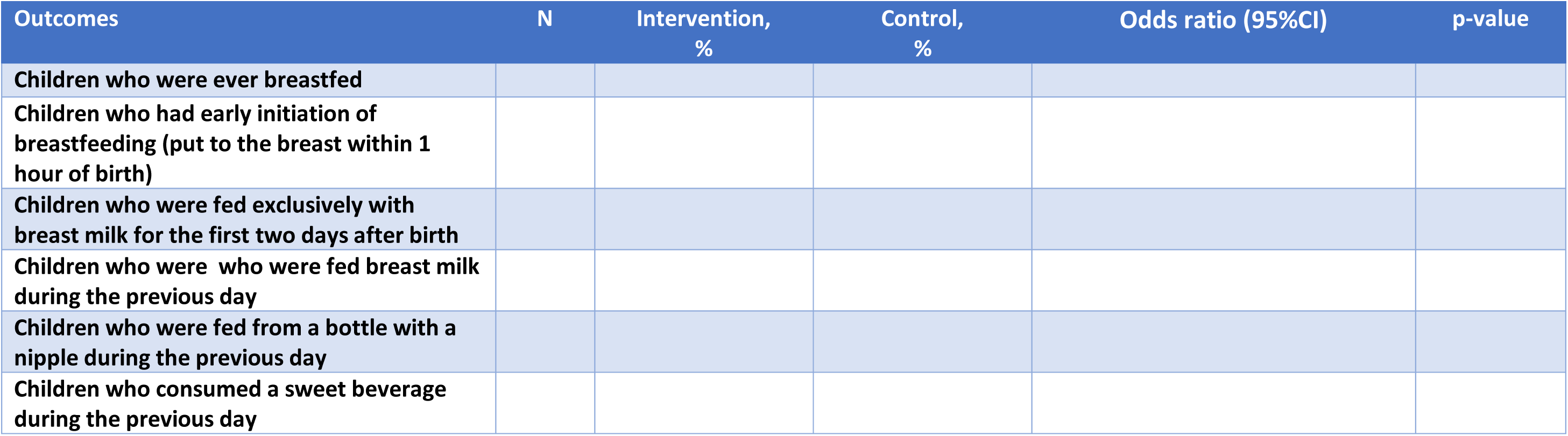
Analyses of IYCF outcomes.

**Figure 1.** Trial profile.

**Figure 2.** Missing data pattern.

**Figure 3.** Distribution of measurements – Hb, SBP, DBP, EQ-VAS, PHQ-9 and GAD-7 scores at each visit, by arm.

**Figure 5.** Forest plot for subgroup analyses.

**Figure 6.** Tipping point analysis.

## Supplementary Tables

**Table A 1.** Adverse pregnancy outcomes.

|  | Intervention | Control |
| --- | --- | --- |
| Miscarriage |  |  |
| Abortion |  |  |
| Still birth |  |  |
| Maternal mortality |  |  |
| Infant mortality |  |  |

**Table A 2.**
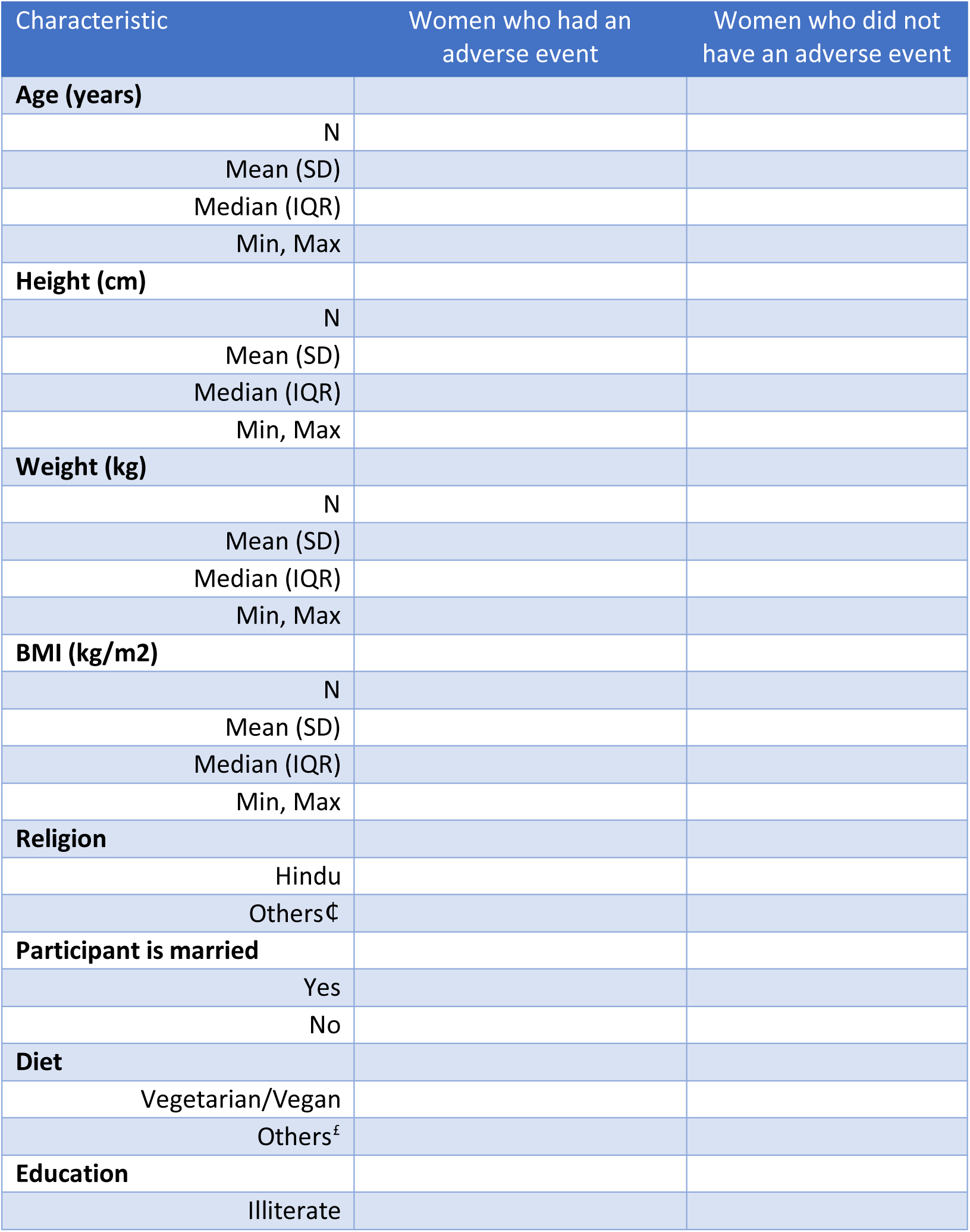
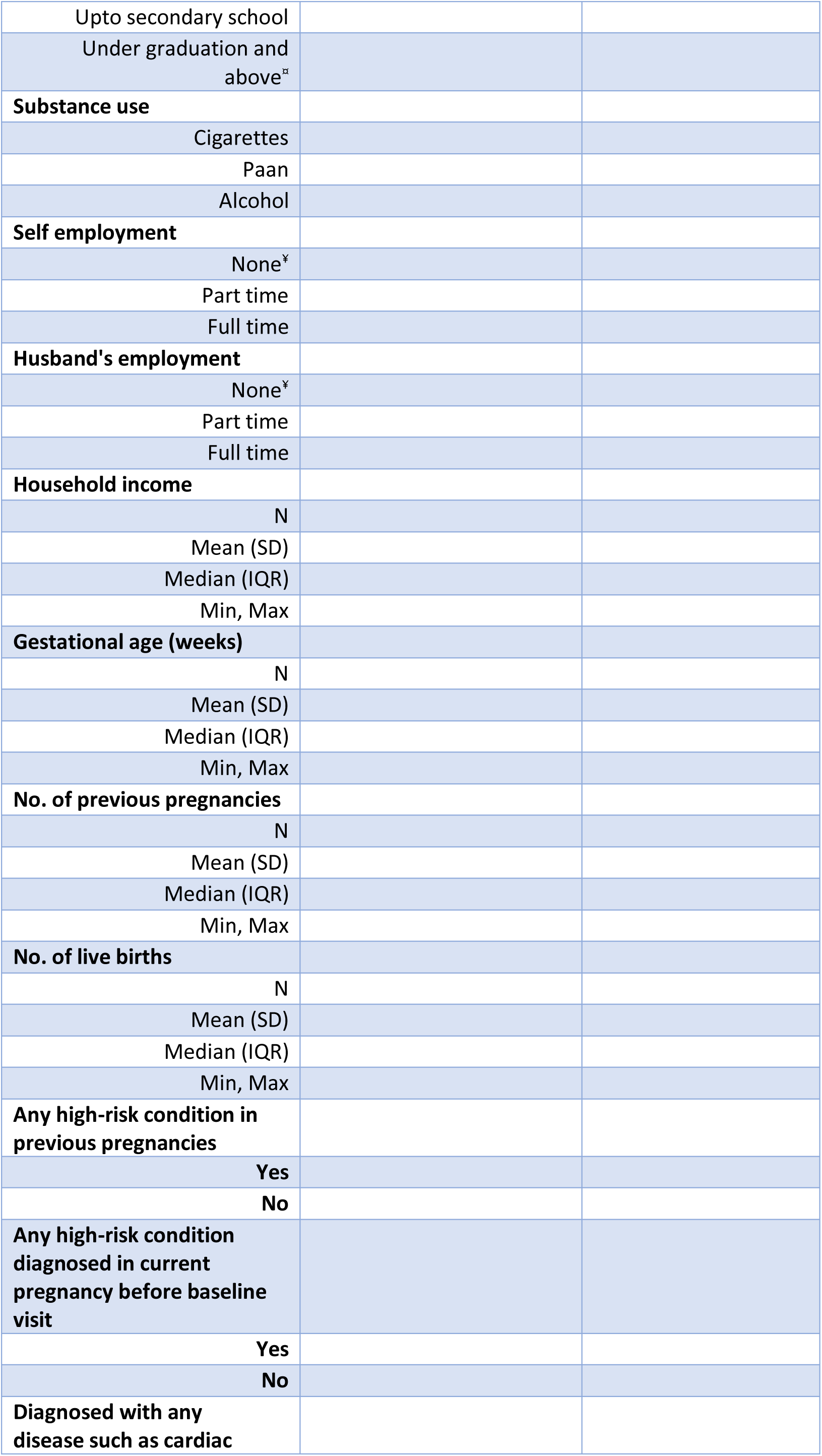
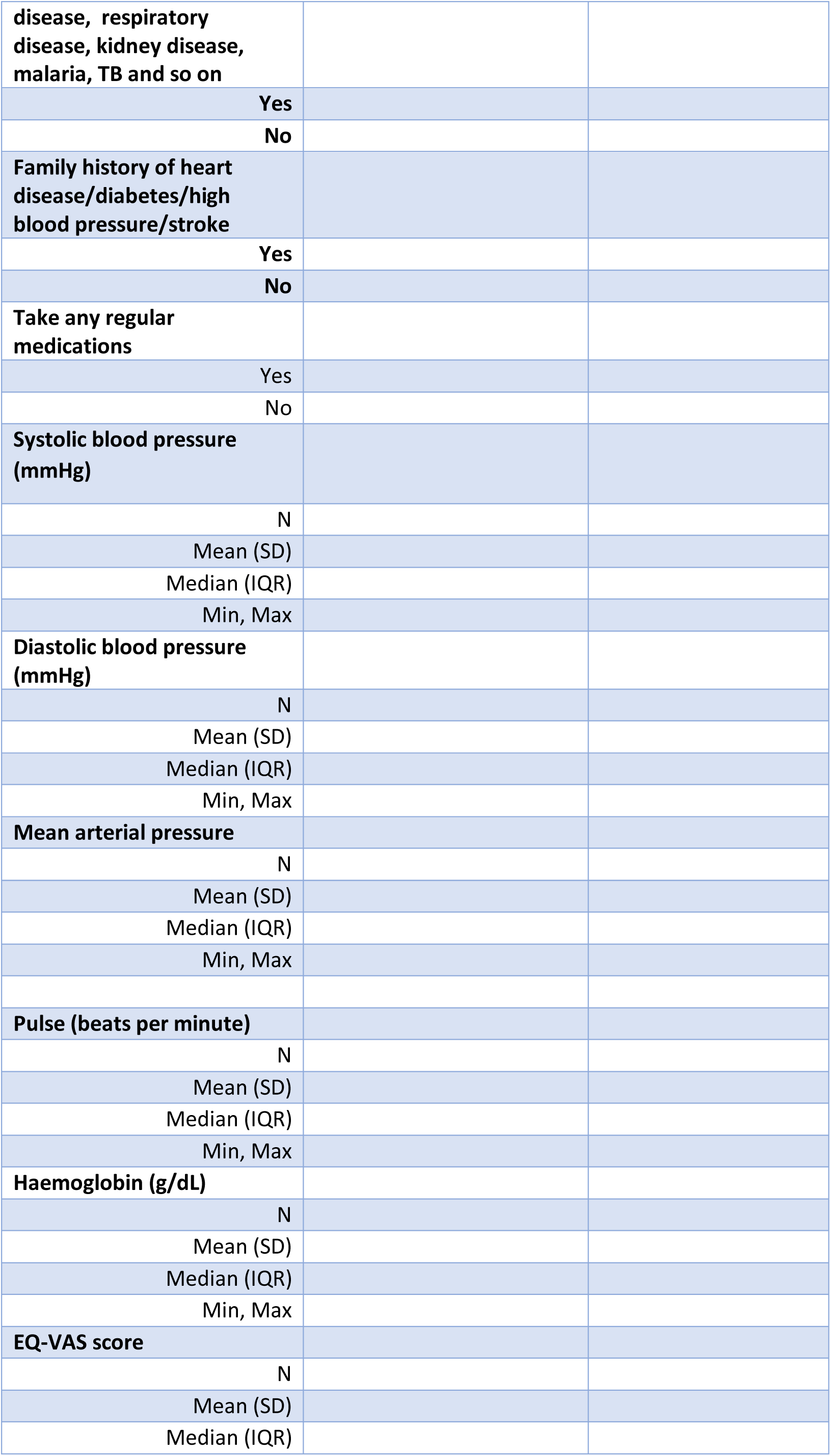
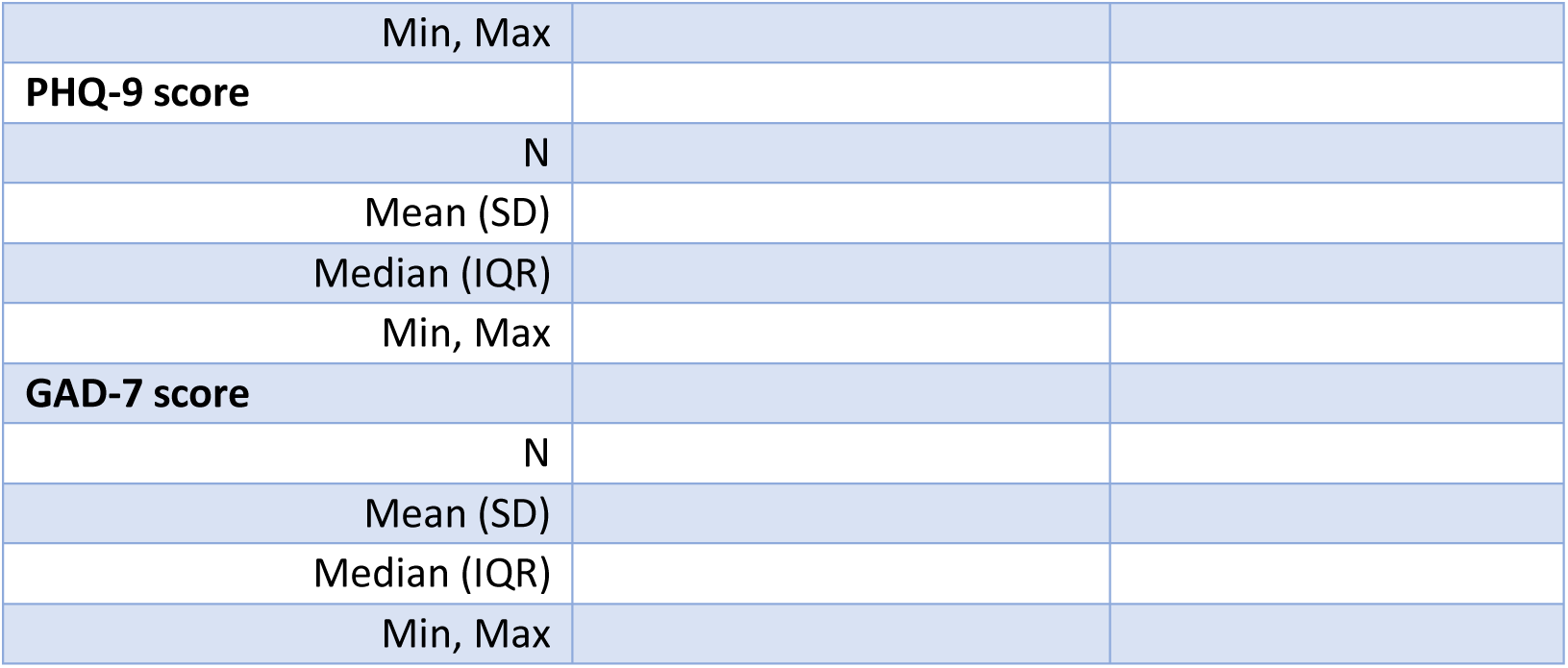
Baseline characteristics of women who had an adverse event vs. those who did not.

**Table A 3.**
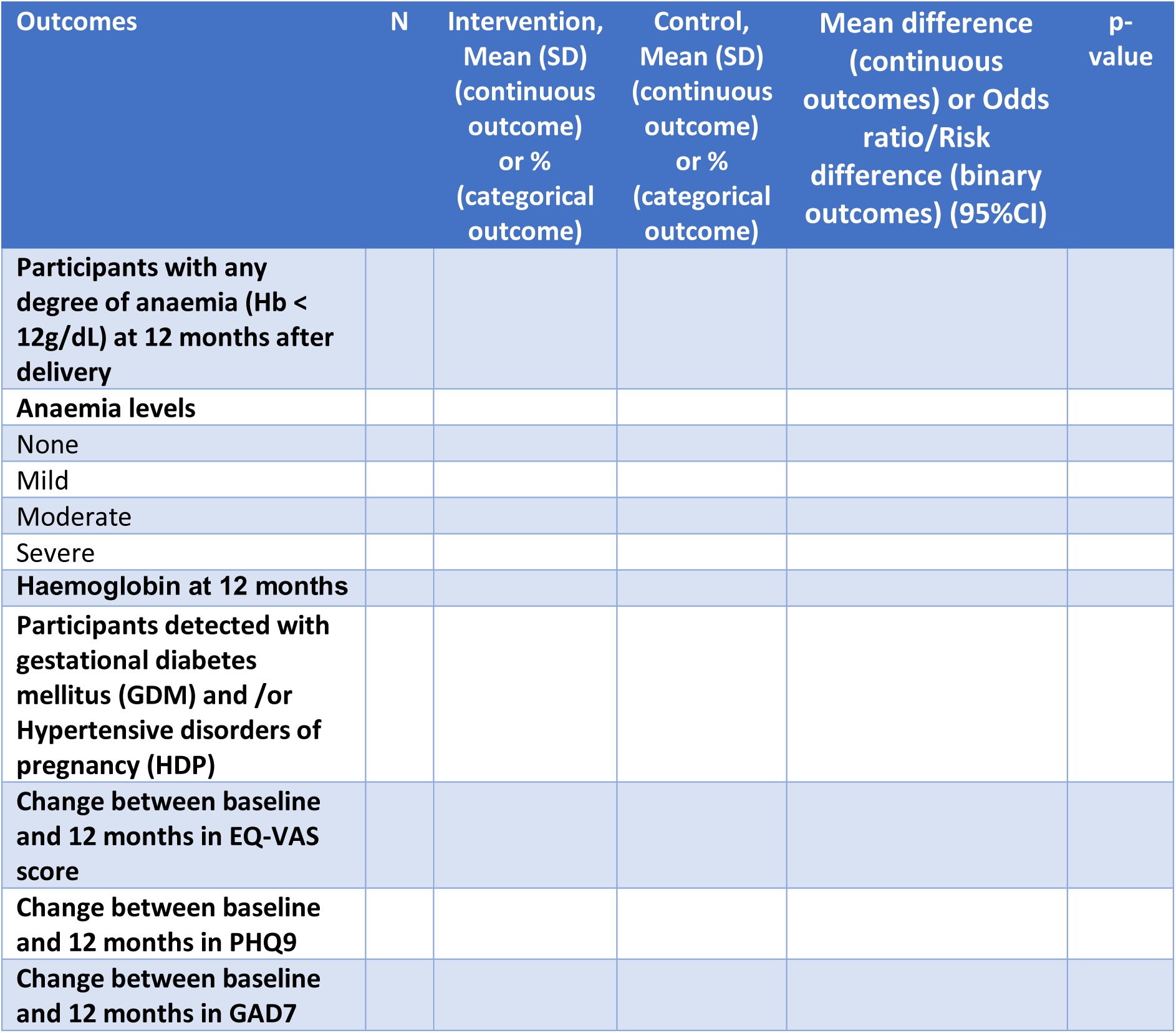
Analyses of primary and secondary outcomes, adjusted for key covariates.

**Table A 4.**
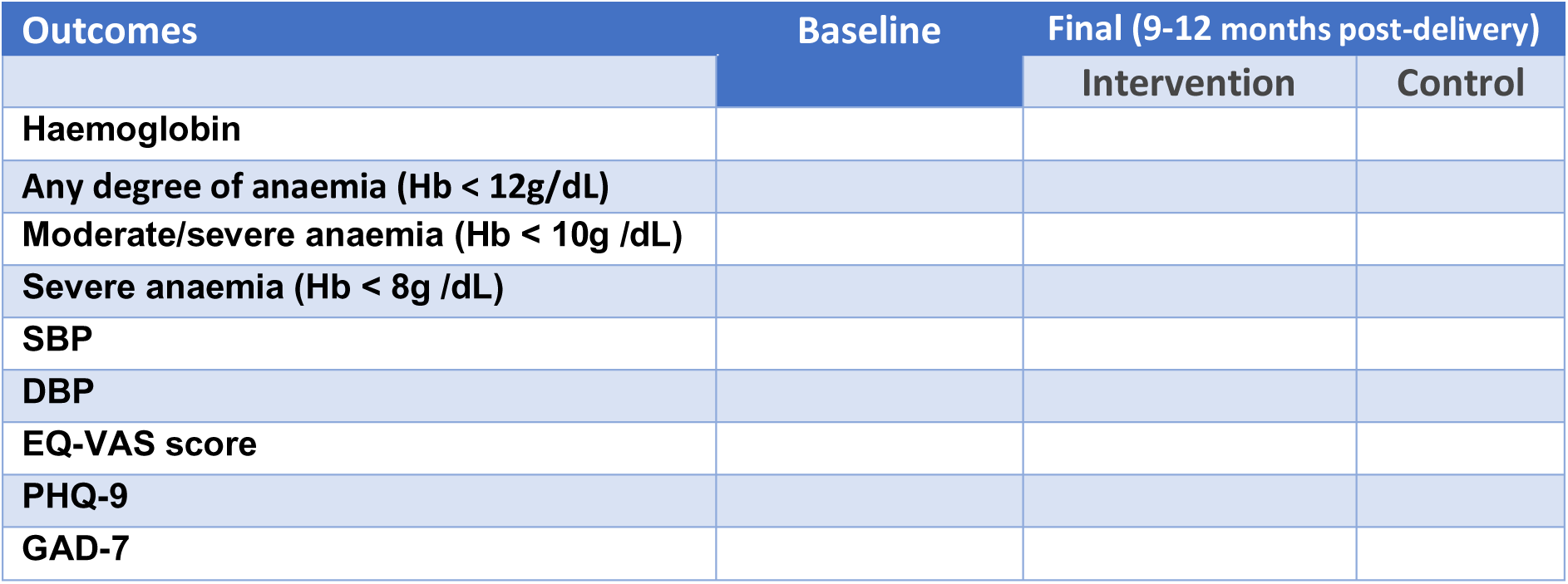
ICC for different outcomes.

**Table A 5.**
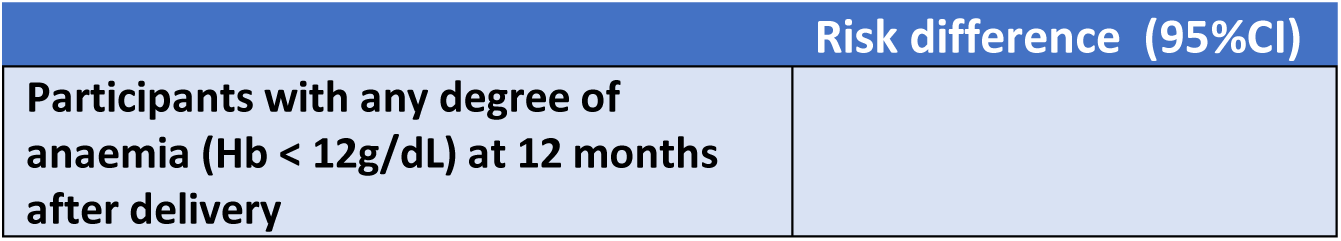
Intervention effect – risk difference along with 95% CI, for anemia (Hb < 12g/dL) at 12 months after delivery.

